# SARS-COV-2 INFECTION IN PRIMARY CARE: A SINGLE-CENTERED, RETROSPECTIVE, OBSERVATIONAL STUDY

**DOI:** 10.1101/2021.11.30.21267074

**Authors:** Pilar Galicia García de Yébenes, Juan José Gutiérrez Cuevas, Fang Fang Chen Chen, Laura Santos Larrégola, Alberto Manzanares Briega, Manuel Linares Rufo

## Abstract

**Purpose:** to describe the clinical characteristics of patients with confirmed SARS-CoV-2 infection in primary care and to analyze the predictive role of different risk factors on prognosis, especially living conditions.

**Methods:** Retrospective longitudinal observational retrospective study by reviewing medical records from a primary care center since March 1 to April 30, 2020. Case definition of confirmed SARS-CoV-2 infection, sociodemographic data, clinical characteristics, comorbidity and living conditions were collected. The statistical analysis consisted in description of the sample, comparison of prognosis groups and analysis of prognostic factors.

**Results:** A sample of 70 patients with confirmed SARS-CoV-2 infection was obtained, with comorbidity mainly related to arterial hypertension, overweight/obesity, hypercholesterolemia, diabetes and chronic pulmonary pathology. Pneumonia was present in 66%. Exitus occurred in 14% of the sample. Factors associated with mortality were advanced age (84 vs 55; p<0.0001), arterial hypertension (78% vs 41%; p=0.040), asthma-COPD (56% vs 13%; p=0.008) and atrial fibrillation (56% vs 5%; p=0.001).

**Conclusions:** The study reflects the clinical practice of a primary care center. This kind of studies are essential to strengthen and reorganize the Health System and to try to anticipate the medium- to long-term consequences of COVID-19 on global health.

## INTRODUCTION

The first cases of atypical pneumonia produced by a new betacoronavirus, SARS-CoV-2 (severe acute respiratory syndrome coronavirus 2), a respiratory virus transmitted by droplets through close contact with an infected person, aerosols, or through fomites in their immediate environment, were described in December 2019 (1-3). A significant proportion of those infected are asymptomatic but can also cause transmission, which constitutes an important challenge for the control of the disease. (4).

The disease, known as COVID-19, has become a pandemic that, as of January 14, 2021, has resulted in more than 90 million people infected and nearly 2 million deaths worldwide (5). The clinical picture is highly variable, with mild involvement in up to 81% of cases, requiring hospital admission in about 14%, and admission to the ICU in the remaining 5% (6). The most common symptoms at the onset of the disease are cough, fever, dyspnea, malaise, anosmia and dysgeusia. In addition, gastrointestinal symptoms, neurological manifestations, hypercoagulability syndrome with venous thrombosis, and skin involvement may occur (4).

The incubation period is 3-6 days. In general, disease progression and the need for hospitalization occur 7-8 days after the onset of symptoms, with rapid deterioration and severe hypoxia indicating the presence of acute respiratory distress syndrome (ARDS). The severity of the disease, characterized by a syndrome of hyperinflammation with cytokine release and potential organ involvement, is mainly related to age and previous comorbidity, especially hypertension, diabetes and cardiovascular disease (7, 8). On the other hand, some biological markers of poor prognosis have been reported, such as lymphopenia and increased of D-dimer, among others (4, 6). Imaging techniques can show from normal radiographs to peripherally distributed bilateral alveolus-interstitial pneumonias (1, 9). Histology shows a significant inflammatory infiltrate characterized by diffuse alveolar damage, capillary congestion, hyaline membrane formation, and diffuse thrombosis of small pulmonary arteries (10). An overall case fatality rate of about 2% has been published (6), although the figures vary greatly depending on the population group considered

As it is an emerging virus, there is no specific treatment available, although different therapeutic options have been used with varying results (11). Consequently, management consists mainly of supportive measures and treatment of symptoms, with anti-inflammatory drugs, such as dexamethasone, and some immunomodulators (12, 13). On the other hand, the prevention of the disease is based on the control of the sources of infection with individual protection measures (personal hygiene and masks) and the study of contacts, as well as on the use of public health measures based on isolation, social distance and community contention (6, 14). In addition, several vaccine prototypes have been developed (15) and are currently being implemented with a population based strategy.

The main publications on COVID-19 have focused on the hospital management of the most severe patients, with little information on the situation of patients in primary care (PC) and the impact of the disease at this level of care. Bearing in mind that PC is the gateway to the system and that 80% of those infected show a mild and self-limited clinical picture, it is essential to know the burden of disease in order to better organize care circuits, and to evaluate possible predictors that can help PC physicians to estimate the risk of hospitalization and possible complications in patients with confirmed SARS-CoV-2 infection.

The purpose of this study is to describe the clinical characteristics of patients with confirmed SARS-CoV-2 infection in primary care and to analyze the predictive role of different risk factors on prognosis, especially living conditions.

## METHODS

### Study design and participants

This is a population-based retrospective and observational study based on data collection from medical records of the population attended by a primary care centre (PCC) in Madrid (Spain). The “Buenos Aires” PCC attends a population of 20,878 inhabitants of the Vallecas area, organized into 12 adult patient quotas of approximately 1,700 patients each one. This is an eminently urban population, with a medium-low socioeconomic level.

The target population consisted of all subjects attended by the center at the beginning of the study. The accessible population consists of all persons with an active clinical history. For the selection of subjects the following inclusion criteria were used: a) over 18 years of age; b) belonging to the reference population of the PCC quotas; c) have been assisted at this PCC, either in person or by telephone, between March 1 and April 30, 2020; and d) have a diagnosis of SARS-CoV-2 infection confirmed by PCR (polymerase chain reaction) during the study period.

### Data collection

We reviewed clinical electronic medical records for all patients with laboratory confirmed SARS-CoV-2 infection. We collected data on sociodemographics (age, gender, country of origin); comorbidity (high blood pressure, hypercholesterolemia, overweight, diabetes, cardiovascular and ictus, renal failure, atrial fibrillation, hypothyroidism, dementia, malignancy, chronic obstructive pulmonary disease (COPD) and smoking); concomitant treatments (ACE inhibitors, steroids, NSAIDs); symptoms of SARS-CoV-2 infection (cough, dyspnea, fever, odynophagia, headache, diarrhea, anosmia, ageusia, skin symptoms); radiological and laboratory findings on admission (leukocyte, lymphocyte and platelet counts, ferritin, creatinkinase, procalcitonin, C-reactive protein, transaminases, D-dimer, pro-BNP), treatment received; living conditions (number of cohabitants, number of bathrooms in the house, elevator, type of transport, and job position, number of times the household was left for shopping or leisure on a daily basis).

### Variables

The outcome variable is poor prognosis, defined by exitus.

### Statistical analysis

The purpose of the study is to describe the clinical characteristics and outcomes of patients. Therefore, there were no formal hypotheses being implemented to drive the sample size calculation and we included the maximum number of patients who met the inclusion criteria.

The description of the sample was expressed as measures of central tendency and dispersion, as well as frequency and percentages for quantitative and qualitative variables, respectively. Differences between exitus and non-exitus were assessed using two-sample t test o Wilcoxon rank-sum test depending on parametric or non-parametric data for continuous variables and Fisher’s exact test for categorical variables. Tests were two-sided with significance set at alpha less than 0.05.

The study was carried out in accordance with the Declaration of Helsinki and the international standards, contained in the International Guidelines for Ethical Review of Epidemiological Studies (Council for the International Organizations of Medical Sciences-CIOMS-Geneva, 1991) and the recommendations of the Spanish Society of Epidemiology (SEE). The study was approved by the local Clinical Research Ethics Committee (Comisión Local de Investigación Sureste). As this is a retrospective study that does not involve a visit with the patient and the person who has access to the clinical history is the physician in charge, the CEICm was asked to waive informed consent for the patients.

## RESULTS

The sample consisted of 70 patients with SARS-CoV-2 infection, confirmed by Ig G or PCR+, with a mean age of 60 ± 21 years, 51.4% of whom were men. Most of them were Spanish (80%) followed by those from Latin American countries (18%). In terms of occupation, most of them were either unemployed or retired (53%), or belong to the socio-health sector (19%). The most frequent comorbidities diagnosed in the clinical record were arterial hypertension (HT) (49%), overweight-obesity (47%), hypercholesterolemia (36%), diabetes mellitus (23%) and chronic pulmonary disease (asthma-CPD) (21%). Twenty-four percent of the patients were taking ACE inhibitors. The most common symptoms were cough (79%), fever (72%), dyspnoea (65%), headache (24%) diarrhea (24%), and less frequently ageusia (9%) and anosmia (8%) (table 1).

**Table 1.** Description of the sample.

| **Characteristics** | **N total** | **n (%)** |
| --- | --- | --- |
| **Sociodemographics** |  |  |
| Men | 70 | 36 (51.4%) |
| Country of origin | 67 |  |
| • Latin |  | 12 (18.2%) |
| • Spain |  | 53 (80.3%) |
| • Romania |  | 1 (1.5%) |
| Occupation | 62 |  |
| • Unemployed or retired |  | 33 (53.3%) |
| • Social-health |  | 12 (19.3%) |
| • Cleaning |  | 3 (4.8%) |
| • Supermarkets |  | 2 (3.2%) |
| • Construction |  | 4 (6.4%) |
| • Industry |  | 3 (4.8%) |
| • Courier/transportation |  | 3 (4.8%) |
| • Security |  | 2 (3.2%) |
| Smoking | 69 |  |
| • No smoker |  | 54 (78.3%) |
| • Ex - smoker (>1 year without smoking) |  | 13 (18.8%) |
| • Current smoker |  | 2 (2.9%) |
| **Comorbidity** |  |  |
| • Hypercholesterolemia | 70 | 25 (35.7%) |
| • High blood pressure | 70 | 34 (48.6%) |
| • Overweight | 68 | 32 (47.1%) |
| • Hypothyroidism | 70 | 12 (17.4%) |
| • Stroke/TIA | 70 | 7 (10.0%) |
| • Ischemic heart disease | 70 | 6 (8.6%) |
| • Asthma-COPD | 70 | 15 (21.4%) |
| • Chronic renal disease | 70 | 2 (2.9%) |
| • Atrial fibrillation | 70 | 8 (11.4%) |
| • Active malignancy | 70 | 2 (2.9%) |
| • Immunosuppression | 70 | 2 (2.8%) |
| • Dementia | 70 | 4 (5.7%) |
| • Diabetes mellitus | 70 | 16 (22.9%) |
| **Concomitant treatments** |  |  |
| • Inmunosupresores | 61 | 1 (1.6%) |
| • ACEs | 70 | 17 (24.3%) |
| • Inhaled steroids | 70 | 6 (8.6%) |
| • NSAIDs | 70 | 5 (7.1%) |
| Age | 60 |  |
| • Mean±SD |  | 59.7±20.6 |
| • Median (P25-P75) |  | 59.1 (45.8-76.0) |
| • Minimum-maximum |  | 18.8 – 93.0 |

Sixty-seven percent of the patients were referred to the hospital while 33% remained under primary care. As for the treatments received, the most common were hydroxychloroquine (74%), azithromycin or ceftriaxone-cefaxone (56%), and low molecular weight heparin (58%). Only 36% received specific antivirals such as lopinavir-ritonavir, with no use of remdesivir. Eleven percent were treated with corticosteroids. As for the analytical parameters, the number of patients for whom this information is available is very variable, although the median values of some of them are presented, such as leukocytes (6485), CRP (41), D-dimer (740), ferritin (510). Of the patients studied, 14% resulted in exitus. In the patients with chest x-ray, 66% had lobar/bilobar pneumonia (16%) or bilateral pneumonia (50%) (Table 2).

**Table 2.**
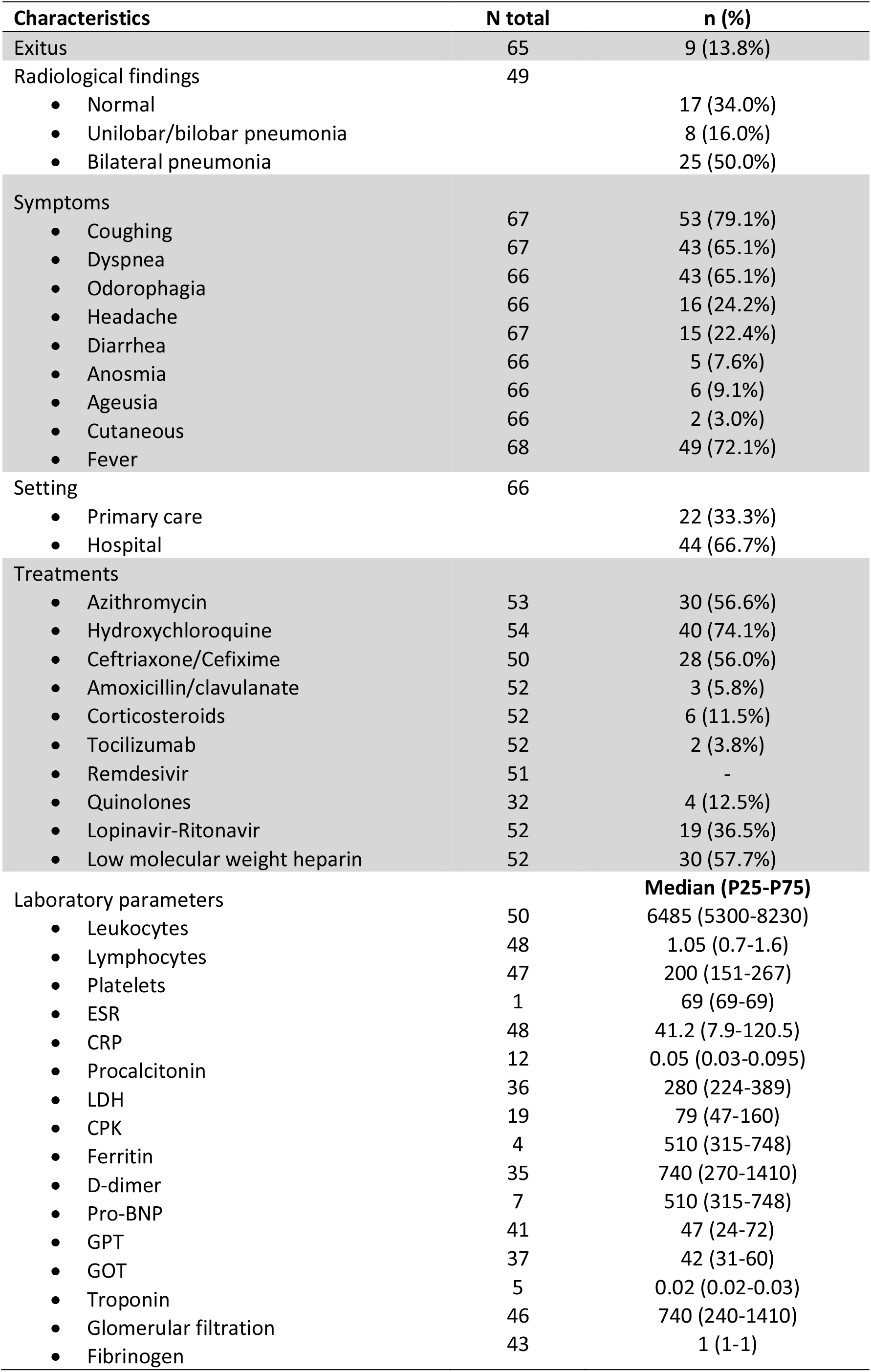
Characteristics related with the SARS-CoV-2.

**Table 3.** Living conditions.

| **Characteristics** | **N total** | **n (%)** |
| --- | --- | --- |
| Number of cohabitans (including the patient) | 54 |  |
| • One-two |  | 38 (70.4%) |
| • Three to seven |  | 14 (25.9) |
| • Twenty |  | 2 (3.7%) |
| Number of bathroom in the household | 36 |  |
| • One |  | 21 (58.3%) |
| • Two or more |  | 15 (41.7%) |
| Elevator | 41 | 73.2% |
| Left home at least once a week (shopping) | 50 | 40 (80%) |
| Left home daily | 50 | 38 (76.0%) |
| Type of transportation used | 48 |  |
| • Private |  | 35 (72.9%) |
| • Public |  | 13 (27.1%) |

Information on living conditions was not available for all patients. It is noteworthy that two patients lived with up to 20 people in the same household. Most household were equipped with just one bathroom (58%) and only 73% had an elevator. Eighty percent of the patients left home at least once a week and 76% went out daily. Twenty-seven percent used public transport.

Among 65 patients with vital status information, 9 resulted in exitus (14%). Patients who died were older (mean age 84 vs 55; p<0.0001) and with higher prevalence of different risk factors as high blood pressure (78% vs 41%; p=0.040), asthma-COPD (56% vs 13%; p=0.008) and atrial fibrillation (56% vs 5%; p=0.001). In addition, the prevalence of overweight (71% vs 41%), stroke (22% vs 9%) and diabetes mellitus (33% vs 20%) was also higher among the exitus, although these differences did not reach statistical significance. On the other hand, no significant differences were observed between exitus and non-exitus in the symptoms of presentation of Covid infection, in the treatments received, or in their living conditions. However, there were differences in derivation, since 100% of exitus occurred in the hospital, probably because they were more severe patients (table 4).

**Table 4.**
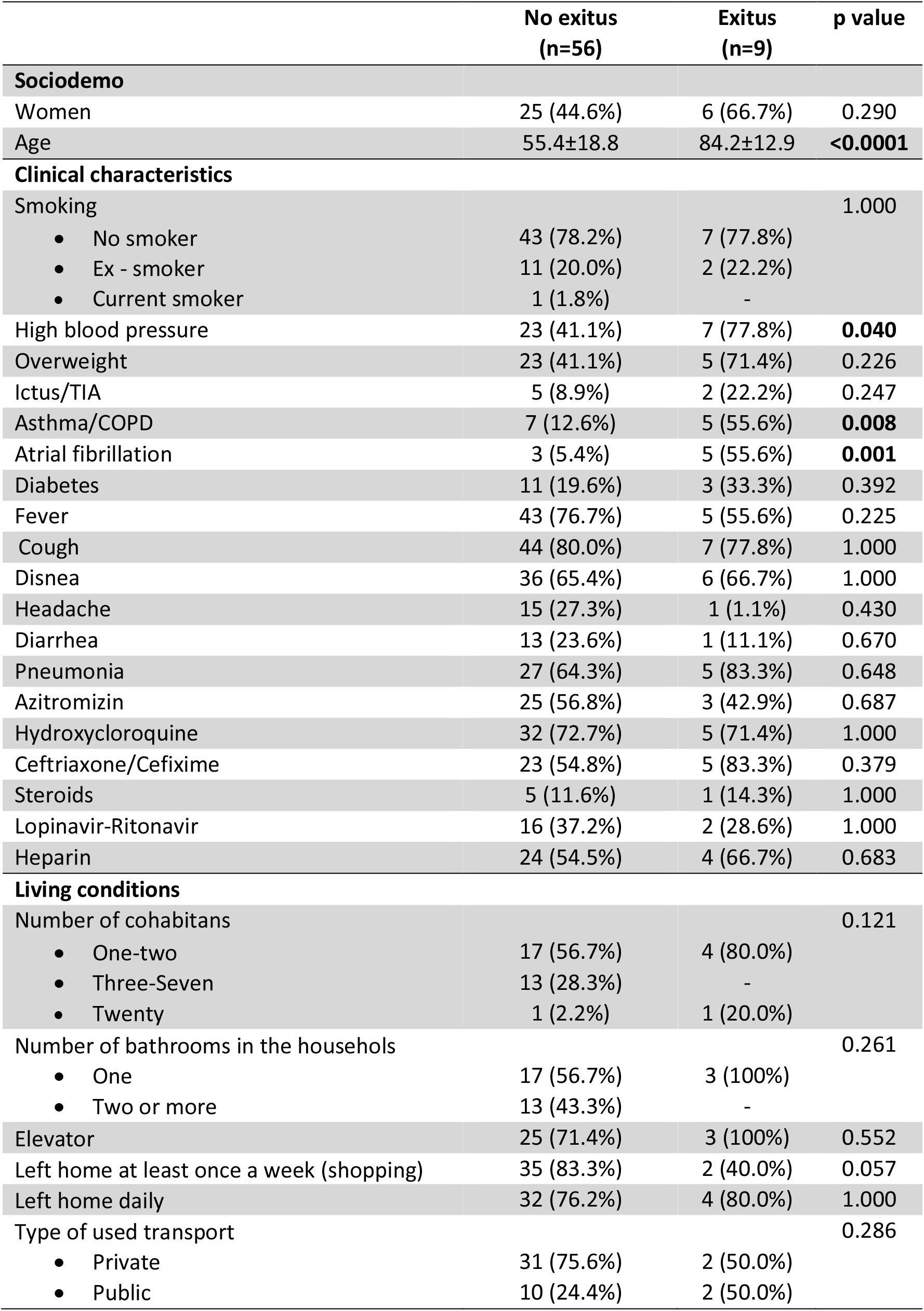
Comparison of groups by prognosis.

The small sample size and the limited availability of information on some variables did not allow us to study the determinants of mortality by multivariate regression analysis in order to estimate the independent effect of possible predictors.

## DISCUSSION

This retrospective population-based study describes the clinical characteristics of SARS-CoV-2 infection in a primary care center in the Madrid region and analyzes possible factors associated with poor prognosis defined as exitus.

The results of this study show an overall mortality of 13.8%, a value in line with literature data. In a meta-analysis of 611,583 subjects from 5 registries in different countries, an overall mortality rate of 12.1% was observed, with wide variations between countries, probably related to the different age distributions (16). In our series, 7 of the 9 exitus were over 80 years of age (77.8%), and only 1 was under 60 years of age (11.1%); in the remaining, age data were not available.

In relation to possible prognostic factors, our data show a higher frequency of exitus in older individuals with arterial hypertension, chronic pulmonary disease, and atrial fibrillation. These types of associations have been confirmed in multiple studies (1, 6, 8, 9, 17) and are explained by the role of the angiotensin-converting enzyme receptor (ACE-2). Many elderly patients with the aforementioned comorbidities are treated with angiotensin-converting enzyme inhibitors (ACEi) or angiotensin II receptor blockers (ARBs), which markedly increase ACE-2 expression.

Although it has been repeated during the pandemic that the COVID-19 virus does not discriminate among patients, it has been shown that the most disadvantaged social groups have a higher frequency of infection and worse prognosis. Several factors explain the greater vulnerability of disadvantaged social groups. Living in small spaces with a high number of cohabitants and without outdoor spaces, precarious jobs without the possibility of teleworking and the use of public transport increase the risk of respiratory infections and reduce the possibility of social distancing. At the same time, poorer access to health services and higher prevalence of chronic diseases worsen the prognosis. In addition to these factors, poor working conditions, low income, and job and financial uncertainty are also known to trigger stress mechanisms that decrease the competence of the immune system and increase susceptibility to infections and the adoption of risky behaviors (18-20).

The PCC “Buenos Aires” belongs to Vallecas, one of the most disadvantaged neighborhoods in Madrid. Due to the characteristics of the population attended at this center, one of our objectives was to analyze the role of living conditions on patient prognosis. We did not obtain results in this regard, although it is true that since this was a retrospective design, the availability of this information in the clinical history is limited.

This study presented has some strengths and is not without limitations. The main strength is its population base. Despite not including secondary care centers, it is very likely that the patients seen in PC centers are sufficiently representative, since it is the first gateway to the system. It is a population-based study and hospital care was not the first option for many patients during the months of highest virus circulation.

The limitations also include problems of representativeness due to the peculiarities of the population of Vallecas, which cannot be extrapolated to other areas of Madrid, and the possible loss of more serious cases that went directly to the hospital. Another limitation of retrospective studies is the possibility of under-registration, which has been particularly striking in the variables on socioeconomic determinants. Finally, the small sample size and the loss of information on some variables have not allowed multivariate analysis of mortality determinants. Despite these limitations, we believe that the study reflects the reality of clinical practice and adequately describes the characteristics and risk factors of patients with COVID-19 infection in the PC setting. Knowledge of the burden of disease is of paramount importance at this level of care as it is the gateway to the system and has a key role in prevention, protection, promotion and treatment of individuals and communities. PC professionals are particularly concerned about the consequences of COVID on health; the decrease in acute consultations or for preventive activities as well as the delay in chronic patients’ management will have a profound impact on the psychological and socioeconomic well-being of populations. This impact is already visible in vulnerable people and will continue in the medium and long term (21, 22). In these times, the holistic view of PC and the need to strengthen and rethink the system in order to increase its efficiency are of particular importance. The availability of studies at this first level is fundamental for this task. Therefore, knowledge of the characteristics and prognostic factors of SARS-CoV-2 infection in the primary care setting may be the first step in improving disease management, reducing the associated burden, and facilitating larger-scale studies of the predictable impact of SARS-CoV-2 infection in the first link of the health care system.

## Data Availability

All data produced in the present study are available upon reasonable request to the authors

## Abbreviations

SARS-CoV-2: severe acute respiratory syndrome coronavirus 2
COVID-19: severe acute respiratory syndrome coronavirus 2
PC: Primary care
PCC: Primary care center
COPD: chronic obstructive pulmonary disease
NSAIDs: Nonsteroidal anti-inflammatory drugs
ACE inhibitors: Angiotensin-converting enzyme inhibitors
PCR: polymerase chain reaction

